# Machine Diagnosis of Chronic Obstructive Pulmonary Disease using a Novel Fast-Response Capnometer

**DOI:** 10.1101/2023.02.22.23286241

**Authors:** Leeran Talker, Daniel Neville, Laura Wiffen, Ahmed B Selim, Matthew Haines, Julian C Carter, Henry Broomfield, Rui Hen Lim, Gabriel Lambert, BRS study team, Scott T Weiss, Gail Hayward, Thomas Brown, Anoop Chauhan, Ameera X Patel

**Author notes:** The following authors contributed equally and share first authorship (Leeran Talker, Daniel Neville). The following authors contributed equally and share senior authorship (Anoop Chauhan, Ameera X Patel).

## Abstract

**Background:** Although currently most widely used in mechanical ventilation and cardiopulmonary resuscitation, features of the carbon dioxide waveform produced through capnometry have been shown to correlate with V/Q mismatch, dead space volume, type of breathing pattern, and small airway obstruction. This study applied feature engineering and machine learning techniques to capnography data collected by the N-Tidal™ device across four clinical studies to build a classifier that could distinguish CO_2_ recordings (capnograms) of patients with COPD from those without COPD.

**Methods:** Capnography data from four longitudinal observational studies (CBRS, GBRS, CBRS2 and ABRS) was analysed from 295 patients, generating a total of 88,186 capnograms. CO_2_ sensor data was processed using Cambridge Respiratory Innovations’ regulated cloud platform, performing real-time geometric analysis on CO_2_ waveforms to generate 82 physiologic features per capnogram. These features were used to train machine learning classifiers to discriminate COPD from ‘non-COPD’ (a group that included healthy participants and those with other cardiorespiratory conditions); model performance was validated on independent test sets.

**Results:** The best machine learning model (XGBoost) performance provided a class-balanced AUROC of 0·968 ± 0·017 and a positive predictive value (PPV) of 0·911 ± 0·028 for a diagnosis of COPD. The waveform features that are most important for driving classification are related to the alpha angle and expiratory plateau regions. These features correlated with spirometry readings, supporting their proposed properties as markers of COPD.

**Conclusion:** The N-Tidal device can be used to accurately diagnose COPD in near-real-time, lending support to future use in a clinical setting.

**Funding:** NIHR (i4i grant), Innovate UK, SBRI Healthcare and Pfizer OpenAir.

## Introduction

Chronic Obstructive Pulmonary Disease (COPD) is a progressive respiratory disease most associated with a smoking history. It is the third most common cause of mortality worldwide [1], causing 3·28 million deaths in 2019 and affecting 212 million people globally [2].

Although no cure currently exists, early diagnosis and treatment are important to improve lung function and quality of life and reduce exacerbations [3]. The clinical standard for COPD diagnosis is spirometry, which relies upon a patient’s ability to exhale forcefully. However, a major disadvantage of spirometry is that it is effort-dependent and thus unreliable and non-specific [4]. In addition, it is effective at detecting latter stages of disease but has lacked reliability in screening for early and asymptomatic cases [5]. This limitation poses challenges in diagnosing a slowly progressing disease such as COPD with a long asymptomatic pre-prodromal phase [6]. It has been estimated that in the UK, only between 9·4% and 22% of those with COPD have been diagnosed [7], in part due to spirometry’s poor precision of only 63% [8].

Capnography is a widely used technique in critical care and anaesthetics. It has been suggested that features of a high-resolution capnogram could be used to identify phys-iologic patterns associated with respiratory diseases such as COPD [9]. Cambridge Respiratory Innovations’ (CRI) N-Tidal™ device has made it possible to measure CO_2_ concentration reliably and accurately via tidal breathing through a mouthpiece, with a greater time resolution than previously possible [10], making the technique an appealing alternative to spirometry.

The objective of this paper was to apply machine learning (ML) techniques to capnography data collected using the N-Tidal device across four clinical studies. The aim was to build a classifier that could distinguish capnograms of patients with COPD from those without COPD using only one breath recording, while maintaining explainability of outputs, with a clear reference to respiratory physiology. This data was analysed to assess the capability of N-Tidal to function as a point-of-care diagnostic tool for COPD.

## Methods

### Participants

It was vital to ensure any diagnostic classifier could distinguish COPD from other conditions that could cause a similar symptom burden at presentation, since many patients undergoing physiological testing for COPD may have an alternative diagnosis. Therefore, the capnograms used in this analysis were collected from four different observational cohort studies, known as GBRS, ABRS, CBRS and CBRS2, that together provided a dataset with appropriate heteroge-nous medical conditions. A summary of these studies with their objectives can be found in Supplementary Information Table 1. They included patients with COPD, asthma, heartfailure, pneumonia, breathing pattern disorder, motor neuron disease, sleep apnoea, bronchiectasis, pulmonary fibrosis, tracheobronchomalacia, as well as healthy participants.

**Table 1.** Demographic information from the four studies and the separate healthy volunteer cohort. Categorical data are given as a number with its percentage of the total (n (%)). Continuous data given as (median (Q1-Q3)). Smoking history was absent for 31 participants and pack years was absent for 58 participants.

|  | COPD<br>(N=80) | Non-COPD<br>(N=215) | Overall<br>(N=295) |
| --- | --- | --- | --- |
| **Age** | 67 (61-73) | 52 (39-64) | 58 (45-67) |
| **Gender (Female)** | 39 (48.8%) | 133 (61.9%) | 176 (58.5%) |
| **BMI (kg/m<sup>2</sup>)** | 25.2 (22.1-30.0) | 28.9 (24.3-34.3) | 27.9 (23.4-33.3) |
| **Smoking History:** |  |  |  |
| Current smoker | 10 (12.0%) | 9 (4.2%) | 19 (6.4%) |
| Ex-smoker | 66 (82.0%) | 71 (36.8%) | 137 (50.7%) |
| Never smoked | 0 (0%) | 113 (58.5%) | 113 (41.9%) |
| **Pack Years (all)** | 40.0 (31.2-58.8) | 0.0 (0.0-4.0) | 1.0 (0.0-20.0) |
| Current smoker | 47.7 (34.5-57.5) | 23.3 (15.0-33.8) | 31.0 (16.5-50.0) |
| Ex-smoker | 40.0 (31.5-58.3) | 5.0 (2.0-10.0) | 12.5 (3.8-38.0) |

In patients with COPD, diagnoses were made according to NICE guidelines with most patients being GOLD stages three or four. Diagnostic criteria used for other conditions, including asthma, are in the supplementary material alongside each protocol’s inclusion and exclusion criteria. In addition to the four studies noted above, capnography data was collected from 27 volunteers without any respiratory disease between December 2015 and January 2022. Though not part of a formal study, these volunteers provided written informed consent and were screened by a medical doctor to ensure they did not have any confounding cardiorespiratory disease or other co-morbidities. All subjects across the four studies gave informed consent, and their data was handled according to all applicable data protection legislations, including the EU/UK General Data Protection Regulation.

Ethical approval was obtained from the South Central - Berkshire Research Ethics Committee (REC) for GBRS and ABRS, the Yorkshire and the Humber REC for CBRS and the West Midlands Solihull REC for CBRS2.

### Procedures

In all studies, capnography data was serially collected using N-Tidal™ device twice daily for varying lengths of time according to each study’s protocol. The N-Tidal device is a CE-marked medical device regulated in the UK and EU, and has been designed to take accurate, reliable recordings of respired pressure of CO_2_ (pCO_2_) directly from the mouth. The N-Tidal device is unique in its ability to accurately measure pCO_2_ from high CO_2_ to background levels, with a fast response time, meaning that quick changes in the geometry of the pCO_2_ waveform are captured.

Patients performed normal tidal breathing through the N-Tidal device for 75 seconds through a mouthpiece in an effort-independent process. CO_2_ was sampled at 10kHz and reported at 50Hz providing a level of resolution not possible with alternative capnometers [10]. A single episode of use (breath recording) produced a single capnogram, with each respiratory cycle (inspiration and expiration) forming a single waveform.

In addition to capnometry data, the following data was also collected for all four studies: basic demographics, spirometry (taken on the same day as N-Tidal data), and medical history. Other clinical and questionnaire data varied across studies (see supplementary material).

### Feature Engineering

For each capnogram collected using a N-Tidal device, raw pCO_2_ data was first denoised, then individual breaths were separated and breath phases segmented before being stored in a secure cloud database (Figure 1). Individual anomalous breaths within a capnogram that could not be processed were identified and excluded from analysis using automated software built into the N-Tidal cloud platform. Reasons for exclusion included: breathing through the nose, water vapour condensation-compromised sensor readings, incomplete breaths, noisy breaths caused by swallowing or coughing into the device, or cardiogenic oscillations which degraded the signal.

**Figure 1.**
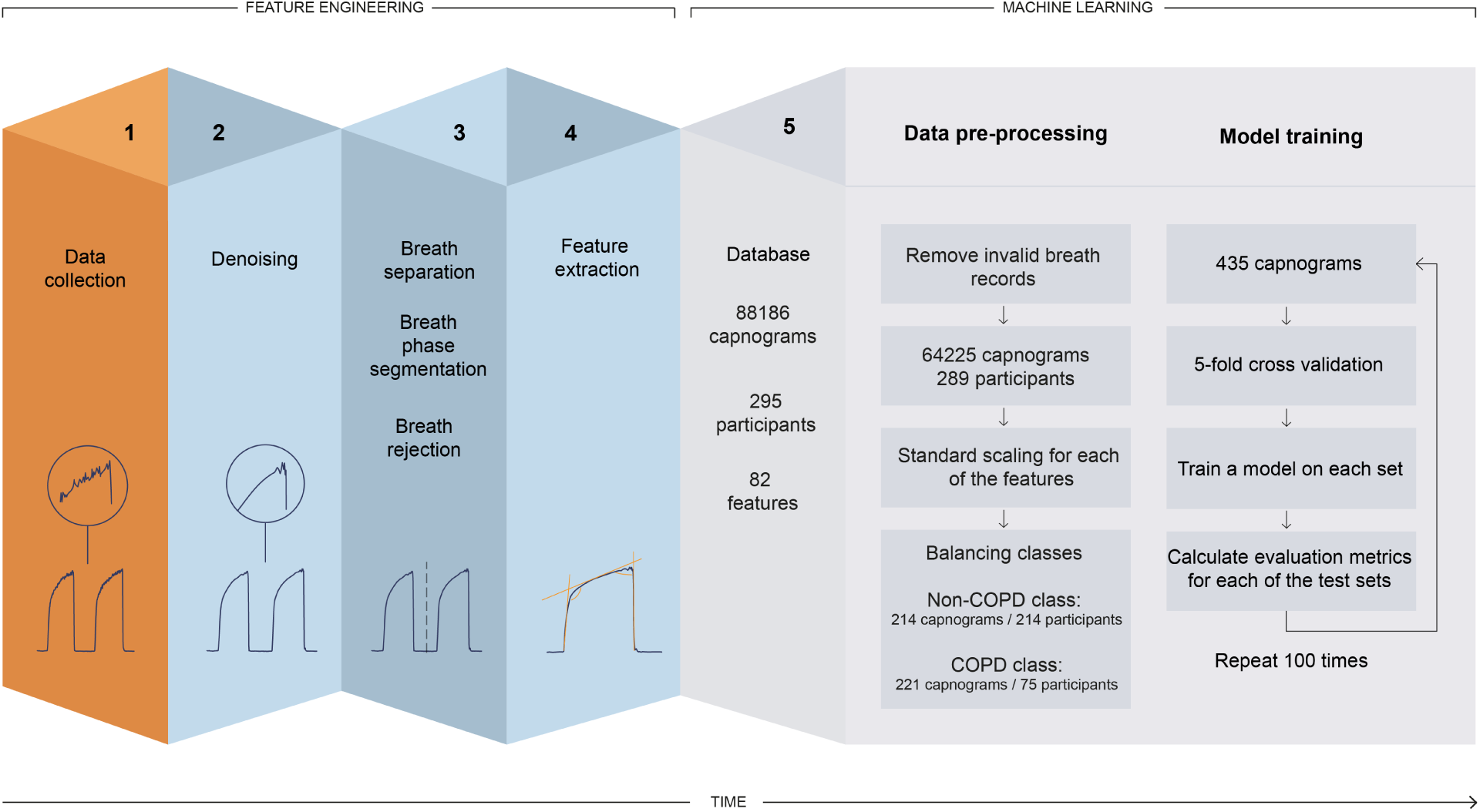
High-level overview of the feature engineering and machine learning pipeline used for data processing, feature engineering, and model training of fast-response CO_2_ data collected through the N-Tidal device.

To generate features for the ML classification task, two categories of information were captured: geometric characteristics of the waveform associated with each breath (referred to as ‘per breath features’); and features of the whole capnogram, such as respiratory rate or maximum end-tidal CO_2_ (ETCO_2_), referred to as ‘whole capnogram features.’

For each capnogram, 77 per-breath features and 5 whole capnogram features were derived. As the number of breaths per capnogram varied, the median and standard deviation were calculated for the per-breath features in each capnogram. These features included the following: *α, β, γ*, and *δ* angles (Figure 2) [11, 12, 13]; gradients and residuals derived from fitting curves to phases, such as the expiratory plateau [14]; absolute and short-term variability of pCO_2_ [15]; curvature and other higher-order time-based features such as the ratio of the expiratory to inspiratory phase [12, 13, 16]; and area under the curve (AUC), which is commonly calculated in volumetric capnography [16, 17]. These features have a basis in the respiratory physiology literature, and in many cases have been hypothesized to relate to clinical airway obstruction.

**Figure 2.**
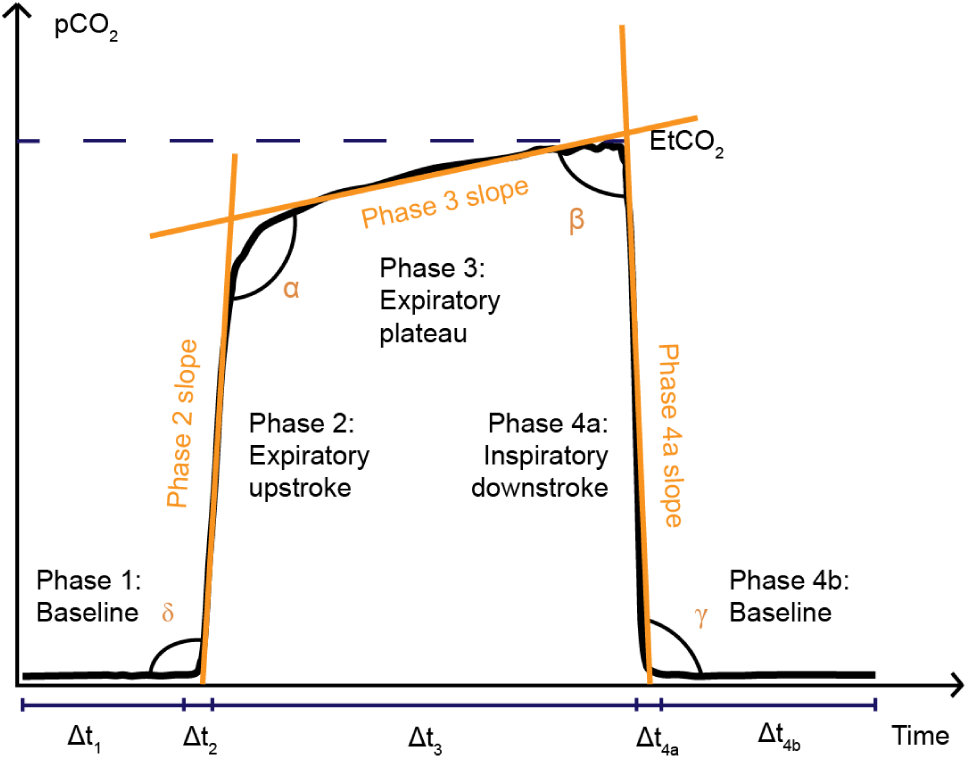
Illustration of a capnogram waveform and its phases and angles. Phase 1 is the inspiratory baseline, Phase 2 is the expiratory upstroke (representing the first p hase o f e xhalation), P hase 3 i s the expiratory plateau (representing the end of exhalation), Phase 4a is the inspiratory downstroke (representing the first phase of inhalation), and Phase 4b is the inspiratory baseline. Note that the start of Phase 1 and the end of Phase 4b may technically be considered part of the same phase.

Any breaths where the full feature set could not be calculated were also automatically excluded from analysis, and further checks were carried out manually to ensure that all condensation-compromised breaths had been excluded by the automated methods.

### Machine Learning

Following pre-processing and feature engineering, each feature was normalized and scaled to a mean of zero and standard deviation of 1. Significant class-imbalances are known to bias ML models towards low predictive accuracy on an under-represented target [18]. To address this, the first capnogram from each participant in the majority class (non-COPD) was retained, and the first three capnograms from each participant in the minority class (COPD) were retained. This approach represented a small fraction of the total breath records available, but using a small number of capnograms per patient had several advantages. First, it ensured that the models would be less likely to overfit on features of individual patients, such as confounding co-morbidities, enabling them to learn the general capnographic characteristics of COPD across a larger population. Secondly, it mimicked the anticipated real-world application of the N-Tidal device and ML model, namely, to diagnose COPD based on a single breath record.

Next, a group-stratified five-fold cross-validation procedure was applied, which involved dividing the dataset into five folds, ensuring no patient overlap between test sets and training sets. For each iteration, data was sampled from four folds for training, and from one retained fold for testing. For example, one iteration of training would use 348 capnograms from 231 patients in the training set, and 87 capnograms from 58 patients in the test set. This ensured that the data used to test performance of the model was not included in the training process. The mean performance of all five iterations produced the results of the overall model performance. In addition, the performance variability across all five folds gave a measure of model generalizability.

The capnogram features were fed to three different ML models: logistic regression (LR); extreme gradient boosted trees (XGBoost); and a support vector machine (SVM) with a linear kernel. These models were chosen due to their more interpretable nature. Deep learning methods were omitted to maintain explainability and to minimize model complexity. The five-fold cross-validation, training, and testing steps were repeated 100 times to calculate the average of the reported metrics. Finally, the capnograms where the model was less confident in assigning to a class (COPD vs. non-COPD) and those that were misclassified were investigated.

### Statistical Methods

The Python scikit-learn package was used to produce each of the following: sensitivity; specificity; negative predictive value (NPV); positive predictive value (PPV); receiver operator characteristic (ROC) curves; micro-averaged area under ROC (AUROC); and confusion matrix (for a decision boundary of 0·5). The most significant features driving model learning were extracted to understand which features of the capnogram waveform were most predictive of COPD. To investigate the relationship between traditional diagnostic methods and these most predictive features, the median of each feature, for each patient, was plotted against the patient’s paired spirometry result, and correlations were calculated.

## Results

Between 6 December 2015 and 31 January 2022, 88186 capnograms were collected from 295 patients. On average, each patient collected 299 capnograms over 179 days. Demographic data was collated (Table 1).

In classifying COPD vs non-COPD participants in the leave out test set, XGBoost showed the best performance with AUROC, specificity and PPV at 0·968, 0·911 and 0·911 respectively (Table 2). SVM showed the best performance in terms of accuracy, sensitivity and NPV at 0·904, 0·901 and 0·899 respectively. Differences were marginal across all metrics, so further analysis is presented for the LR model, as it is the most explainable. These metrics and those in Figure 3 (including standard deviations across folds) are an average across the 100 iterations. In other words, five-fold cross validation was performed 100 times, and the results were averaged across all iterations.

**Table 2.** Machine learning model performance on the leave out test set, averaged across 100 iterations, for each of the three models built: logistic regression (LR), extreme gradient boosted trees (XGBoost), and support vector machine (SVM) with a linear kernel. The highest performance (across all models), for each of the metrics reported is highlighted in bold.

|  | Accuracy | AUROC | Sensitivity | Specificity | Negative Predictive Value (NPV) | Positive Predictive Value (PPV) |
| --- | --- | --- | --- | --- | --- | --- |
| LR | 0.898 ± 0.020 | 0.962 ± 0.013 | 0.888 ± 0.037 | 0.908 ± 0.026 | 0.887 ± 0.034 | 0.908 ± 0.024 |
| XGBoost | 0.900 ± 0.025 | **0.968 ± 0.017** | 0.889 ± 0.040 | **0.911 ± 0.032** | 0.888 ± 0.038 | **0.911 ± 0.028** |
| SVM | **0.904 ± 0.020** | 0.957 ± 0.019 | **0.901 ± 0.038** | 0.908 ± 0.025 | **0.899 ± 0.029** | 0.910 ± 0.023 |

**Figure 3.**
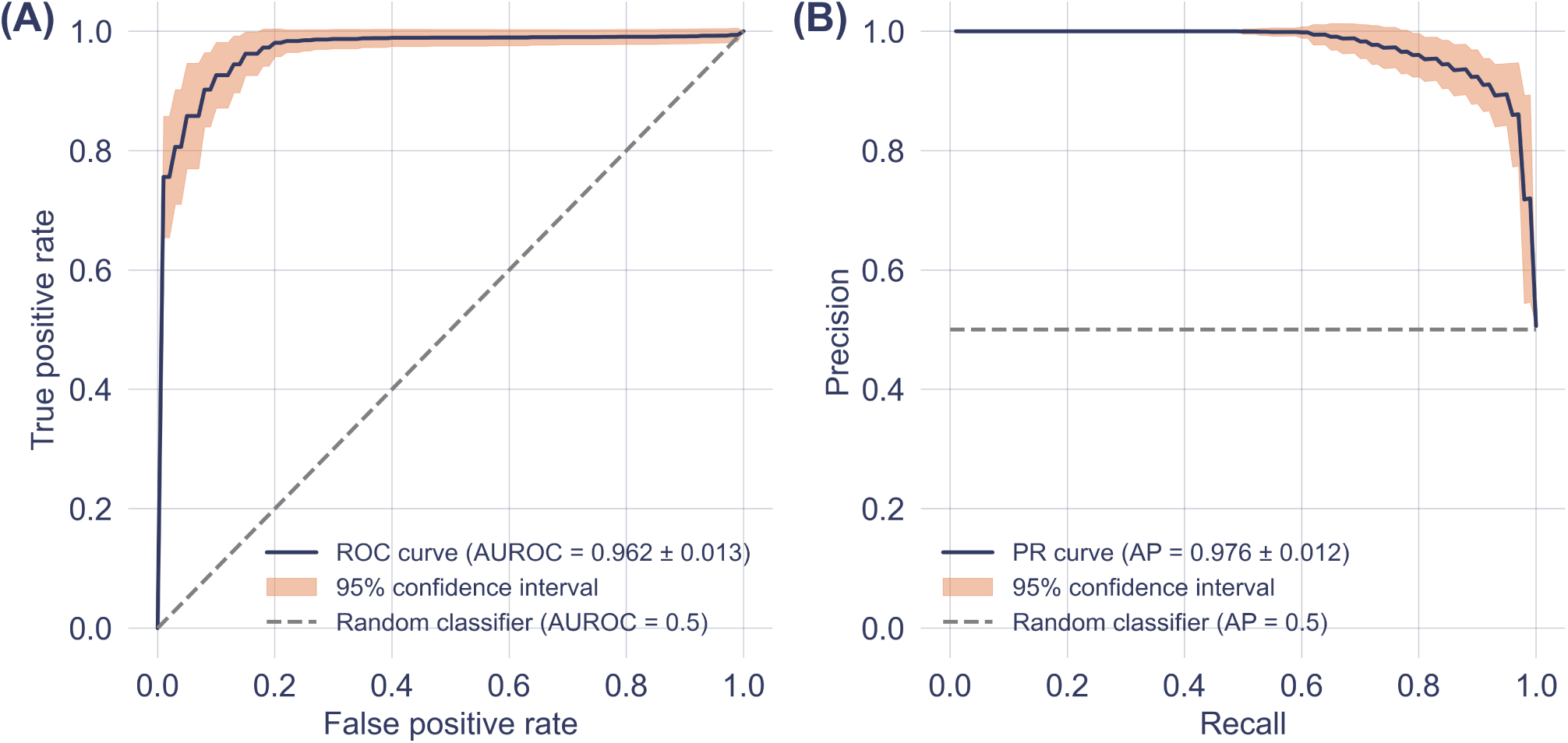
(A) Receiver operating characteristic (ROC) curve for the LR model, reported with results of a theoretical ‘random’ classifier with no predictive power, and a 95% confidence interval, calculated across all 100 iterations. (B) Precision-Recall Curve for the LR model, reported with the results of a theoretical ‘random classifier’ and the average precision (AP).

A post-hoc evaluation of the predictive model was conducted to identify which capnogram features best distinguished patients with and without COPD. The relative feature importances for driving learning in the LR model were determined and the region on the capnogram waveform from which features were extracted was used to construct the importance map for non-COPD and COPD waveforms in Figure 4. The heatmap value of each region represents an average of the weighted feature importance for that region, across all features assigned to that region. The weighted feature importance is calculated as the normalized value for that feature multiplied by the feature importance. Features associated with phase 2 and phase 3 of the capnogram waveform (the exhalation phase) were found to be the most important drivers of learning. Figure 5 shows the average waveform across a 75 second capnogram of four correctly classified capnograms: two each for low and high model confidence, where confidences were calculated as the mean across 100 iterations.

**Figure 4.**
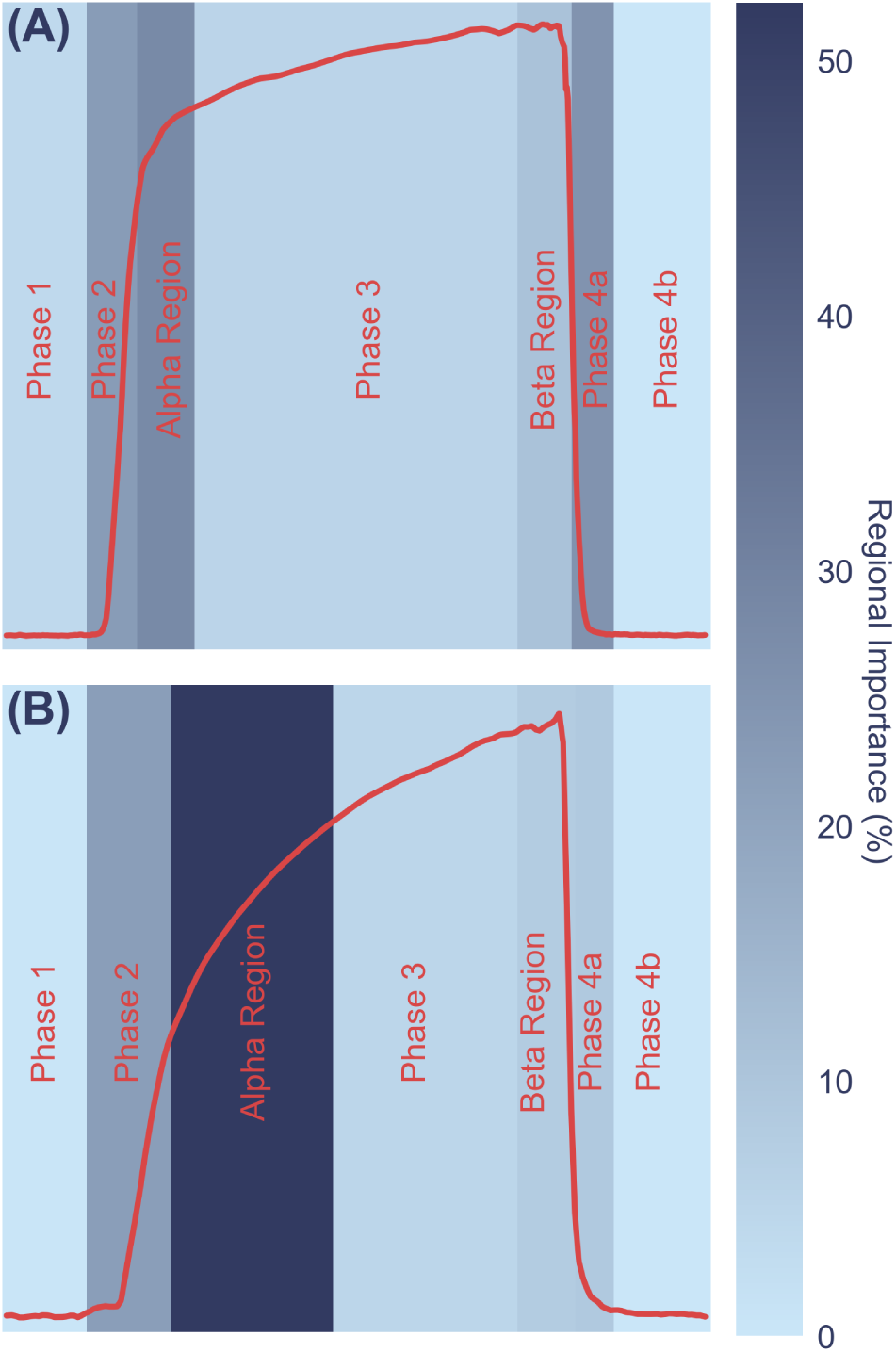
Average weighted feature importance by capnogram waveform region, where weighted features were calculated as the magnitude of the product of the normalized feature value and the feature importance. (A) shows an example for a non-COPD waveform, and (B) shows an example for a COPD waveform.

**Figure 5.**
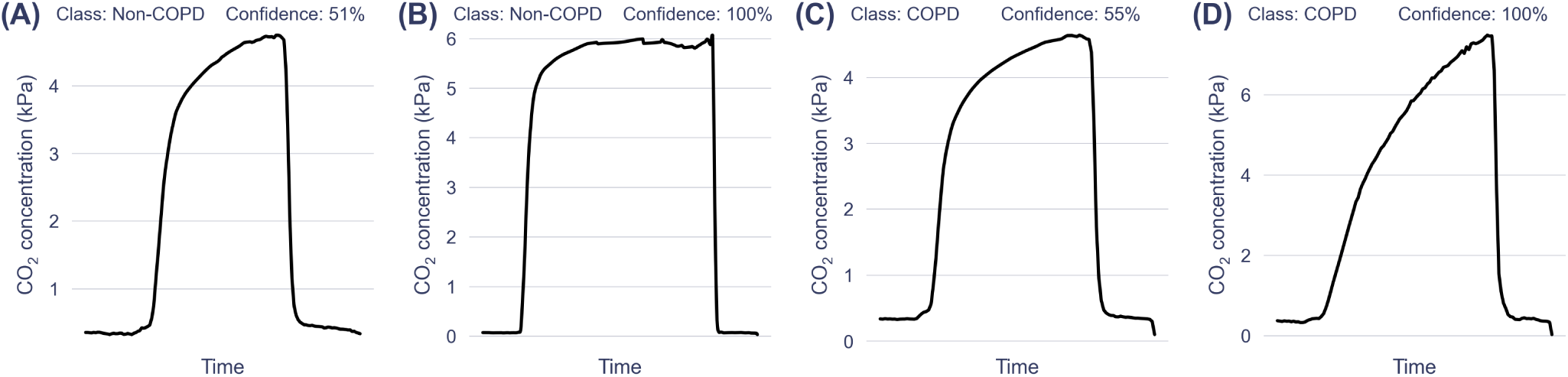
Average waveforms (of correctly classified capnograms), with prediction confidence expressed as a % (averaged across 100 iterations). A and B are two examples of the non-COPD class with the corresponding confidences and C and D show two examples of the COPD class with the corresponding confidences. A and C are examples with low prediction confidence and B and D are examples with high prediction confidence.

To understand the relationship between the capnography features driving the learning and spirometry metrics typically reported in COPD, the FEV_1_ % predicted for each participant was correlated with the corresponding 20 most important features as determined by the LR model, using a Spearman’s rank correlation coefficient, *ρ* (since not all relationships were linear). This was only calculated for patients with paired spirometry (177 patients). Of the 20 features, six showed significant correlations |*ρ*| >0·5. An example of one of the most important feature’s relationships was modelled and plotted against FEV_1_ % predicted (Figure 6), where a linear relationship prompted the calculation of the product moment correlation coefficient (*r*) to be 0·718.

**Figure 6.**
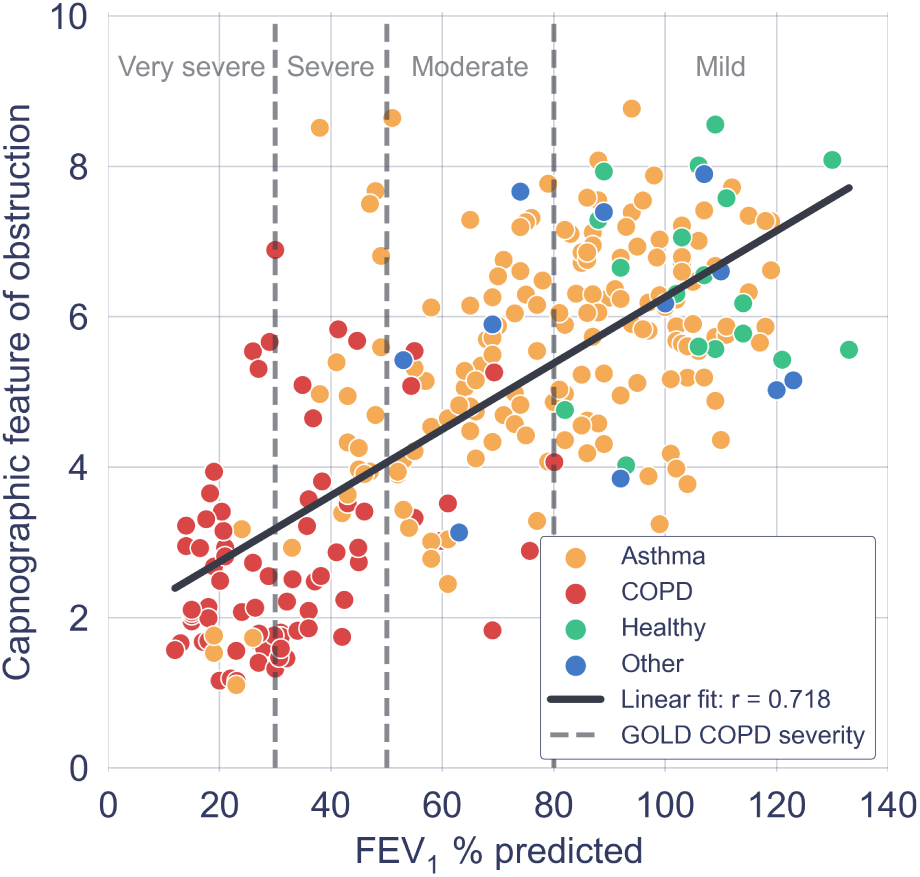
Scatter plot of an example of one of the most important capnography features driving learning in the logistic regression model, versus FEV1 % predicted from paired spirometry data. Each point represents a single paired capnogram. The correlation coefficient was 0.718.

Next, to assess whether there was significant variation in classification results between repeat capnography readings taken over time, the classification accuracy and the standard deviation of the LR model’s output probabilities was calculated for all of the capnograms of each patient in the test fold (as opposed to evaluating only one or three capno-capnograms was not statistically significant (Mann-Whitney U = 8000, P = 0·92 two-tailed), indicating that age did not bias the chances of misclassification. The same procedure was performed for BMI, yielding similar outcomes (Mann-Whitney U = 7700, P = 0·57, two-tailed) gram(s) per patient). The results were averaged over 100 iterations to ensure the consistency of the result, and the results reported in Table 3 are mean values calculated over all the patients in each disease category for both metrics.

**Table 3.** Classification accuracy and standard deviation of the model confidence, by disease group, for all capnography data collected by each participant in the test set, averaged over disease group and again over all 100 iterations.

|  | Number of patients | Average number of capnograms per patient | Classification accuracy per patient | Std. of probability of COPD per patient |
| --- | --- | --- | --- | --- |
| Healthy | 34 | 242 | 0.968 ± 0.006 | 0.072 ± 0.007 |
| Asthma | 141 | 289 | 0.923 ± 0.005 | 0.073 ± 0.002 |
| COPD | 75 | 150 | 0.891 ± 0.009 | 0.101 ± 0.004 |
| Heart Failure | 10 | 169 | 0.798 ± 0.025 | 0.142 ± 0.011 |
| Breathing Pattern Disorder | 10 | 124 | 0.981 ± 0.003 | 0.086 ± 0.006 |
| Pneumonia | 16 | 46 | 0.839 ± 0.016 | 0.105 ± 0.007 |
| Motor Neuron Disease | 3 | 95 | 0.800 ± 0.047 | 0.129 ± 0.016 |

An important aspect of model validation was to investigate demographic bias. For this, patients that were misclassified more than 50% of the time across all 100 iterations were termed ‘misclassified’. These misclassification rates were stratified by birth sex and COPD status, highlighting that discrepancies were small and misclassification was not overly skewed towards a single characteristic (Table 4). Additionally, the difference between the medians of age distributions of the misclassified and correctly classified capnograms was not statistically significant (Mann-Whitney U = 8000, P = 0·92 two-tailed), indicating that age did not bias the chances of misclassification. The same procedure was performed for BMI, yielding similar outcomes (Mann-Whitney U = 7700, P = 0·57, two-tailed).

**Table 4.** Misclassification rates of each sex versus disease group for the logistic regression (LR) model.

|  | Female | Male | Total |
| --- | --- | --- | --- |
| Non-COPD | 7.6% (10/132) | 11.0% (9/82) | 8.9% (19/214) |
| COPD | 8.3% (3/36) | 7.7% (3/39) | 8.0% (6/75) |
| Total | 7.7% (13/168) | 9.9% (12/121) | 8.7% (25/289) |

## Discussion

The aim of this study was to demonstrate that a single breath recording from the N-Tidal™ fast-response capnometer, in conjunction with a machine-learning-derived diagnostic classifier, could be used to accurately classify those patients with and without COPD. It was found that the performance of the best machine learning model (XG- Boost) provided a class balanced AUROC of 0·968 ± 0·017 and positive predictive value (PPV) of 0·911 ± 0·028. The performances of the three models (LR, XGBoost and SVM with a linear kernel) were very similar (Table 2). For instance, the largest difference in sensitivity between models was 0·013 and the largest difference in positive predictive value between models was 0·003. This model-independence provides confidence in the thesis that COPD can be reliably detected from geometric properties of the CO_2_ waveform. Furthermore, the prediction accuracy by disease state implies that healthy individuals and patients with asthma or COPD are the easiest to classify (healthy being the easiest), with 0·968 ± 0·006, 0·923 ± 0·005 and 0·891 ± 0·009 classification accuracies respectively (Table 3). The more consistent classification results for healthy individuals may reflect a reduced variability in CO_2_ waveform shape over time, compared to individuals with an obstructive airway disease with an element of airway variability.

The motivation for using XGBoost, LR and SVM with a linear kernel was to achieve a high level of interpretability, and ease of traceability to the individual features that most informed the model’s decision. The identified features (Figure 4) contributing most to the model’s decision were from the *α* angle region and expiratory plateau, where the latter features quantified the concavity, height, and central timestamp of the expiratory plateau. Together, these features characterize the rate at which gas from the upper airways (poor in CO_2_) gives way to mixed alveolar gas from the lower airways (richer in CO_2_). A larger *α* angle, and a smaller value of these plateau features corresponds to greater concavity and greater airway resistance, likely due to an obstructed bronchospastic airway. The analysis supports the physiological plausibility of these features in underpinning the obstructive airway pathology of COPD, as a number of the most important features showed strong correlation to paired FEV_1_ % predicted data (Figure 6).

The average waveforms of a selection of capnograms for which the model had varying predictive confidence can be seen in Figure 5. Capnograms for which the model had the highest confidence were at the extremities of a square-shaped healthy waveform with smaller *α* angle and smaller expiratory plateau tangent (relative to the horizontal), or a shark-fin-shaped COPD waveform with larger *α* angle and larger expiratory plateau tangent (relative to the horizontal). Reassuringly, we did not find any significant association between prediction accuracy and demographic features (including age, birth sex, and BMI) through non-parametric statistical testing, indicating that there is no systematic bias in model performance with respect to demographic data collected on the studies.

Several factors may have contributed to the erroneous classification of a small group of patients as having COPD when they had asthma or other respiratory conditions. First, smokers and ex-smokers with a significant pack year history were present in the non-COPD group, suggesting there may be some patients with undiagnosed COPD. Of the 25 patients misclassified by the LR model, 12 were either current smokers or ex-smokers, and this group had a mean smoking pack year of 22. Second, the ABRS study highlighted a subgroup of asthma patients whose capnometry was ‘COPD-like’; some patients in this subgroup were suspected to have airway remodelling through many years of severe, poorly controlled asthma, creating a COPD-like clinical picture.

The work presented in this article has a number of limitations. First, simpler ML models were used in keeping with the National Health Service Artificial Intelligence recommendations regarding algorithmic explainability [19]. It is important to understand the link between the features driving the ML model and the pathophysiological process under investigation, but this precludes ‘deep learning’ methods that may have enhanced model performance. Second, COPD patients in the source dataset were predominantly GOLD stages 3 or 4 and managed in secondary care and, therefore, not representative of the general COPD population. Ideally, a diagnostic model would be based on a prospective analysis of those with a clinical suspicion of COPD divided into those who did and did not ultimately have the condition. Third, ground-truth labels could only be obtained using current diagnostic pathways, known to have their shortcomings and inaccuracies. Therefore, a misclassification of a participant by one of the models presented in this article could, in some cases, be caused by mislabelling or misdiagnosis. Finally, the variability in pathologies in the non-COPD group was limited to the datasets collected on the studies, and may fail to account for the full spectrum of heterogeneity that would be encountered in a real-world clinical setting. Regardless, the proposed methods managed to distinguish on capnography alone (without supplementary data such as smoking history), between COPD and a range of differential diagnoses for patents likely to be referred for respiratory physiological testing (including healthy volunteers), demonstrating its potential clinical benefit.

In summary, we demonstrate that the N-Tidal fast-response capnometer and cloud analytics pipeline can perform real-time geometric waveform analysis and machinelearning-based classification to accurately diagnose COPD within minutes of breathing into the device. In contrast to commonly used ‘black box’ machine learning methodologies, a set of highly explainable methods were used that can provide traceability for machine diagnosis back to individual geometric features of the pCO_2_ waveform and their associated physiological properties suggestive of obstructive airways disease.

## Data Availability

A summary of study data is contained in the manuscript; study objectives, inclusion and exclusion criteria are provided in the Supplementary Information. Raw study data is not publicly available due to restrictions by privacy and data protection laws.

## Acknowledgements

The ABRS study was supported by the National Institute for Health Research Invention for Innovation (NIHR i4i) Programme (Grant Reference Number: II-LA-1117-20002), the GBRS study was supported by Innovate UK (Grant Reference Number: 102977), the CBRS study was supported by SBRI Healthcare, and the CBRS2 study was supported by Pfizer OpenAir. The ABRS and GBRS research was supported by the Portsmouth Technology Trials Unit (www.pttu.org.uk) and the NIHR Community Healthcare MedTech and In Vitro Diagnostics Co-Operative (MIC). The authors also acknowledge the work of staff at the Cambridge COPD Centre for Respiratory Research for their role in patient recruitment for CRBS and CBRS2, and staff in Oxford University’s Primary Care Clinical Trials Unit who were responsible for trial management (Julie Allen, Johanna Cook, Joy Rahman, Rebecca Edeson) and nursing (Heather Rutter, Karen Madronal, Bernadette Mundy) in ABRS.

## Author contributions

AC, TB, GH, DN, LW, and the BRS study team conceived of the individual studies, and were responsible for study supervision and study design. DN, LW, and the BRS study team collected the data. MH and AXP conceived of the original models, analyses and experimentation presented in this article. MH, JCC, and AXP designed and wrote the algorithms and code for denoising, pre-processing, and feature engineering. LT, ABS, MH, HB, RHL, AXP wrote the code for, and carried out, model training and data analysis. AXP supervised the analyses in the manuscript. STW, AXP, GL, AC, TB, and GH contributed to expert technical review and data interpretation. LT, GL, ABS, HB, RHL drafted the manuscript. All authors participated in review and editing of the manuscript. All authors had access to the results data presented in the manuscript and all authors read and approved the manuscript. Access to ABRS study data was limited to LT, ABS, MH, JCC, HB, RHL, AXP, LW, TB, AC, GH. Access to the GBRS study data was limited to LT, ABS, MH, JCC, HB, RHL, AXP, DN, TB, AC. Access to CBRS and CBRS2 study data was limited to LT, ABS, MH, JCC, HB, RHL, AXP, and some members of the BRS study team. Restrictions to access of raw data was for data protection reasons. The decision to submit this manuscript was made by all authors. LT and DN contributed equally to this work and share first authorship. AC and AXP contributed equally to this work and share senior authorship.

