## supplementary material for "Machine Diagnosis of Chronic Obstructive Pulmonary Disease using a Novel Fast-Response Capnometer"

---

### Further Study Details

#### COPD Breathing Record Study (CBRS)

**Primary Objective**
To collect a longitudinal observation study database of capnograph records for up to 30 patients with COPD over 6 weeks using the N-Tidal C data-collector capnometer.

**Secondary Objectives**

- To identify the correlation of carbon dioxide ($CO_2$) measurements by capnography with those obtained by periodic standard arterial and capillary blood gas measurements.
- To identify the within-day and day-to-day variability of exhaled carbon dioxide in patients with COPD.
- To measure the absolute change in exhaled carbon dioxide measurements of patients admitted with an acute exacerbation of COPD during their recovery.
- To capture the carbon dioxide signature predictive of exacerbation in monitored patients who undergo an acute admission during the study.

**Device Specific Objectives**

- To assess the ability of patients with COPD to use the N-Tidal C data-collector capnometer daily and to capture user feedback.
- To assess the frequency of replacement of the consumable breath tubes and mouthpieces in normal daily use.

**Inclusion criteria**

- Aged 18 years and over.
- Case managed group only: Diagnosis of COPD; Chronically elevated $PaCO_2$; Susceptible to frequent exacerbations of COPD

- Acute admission group only: Hospitalized via the emergency room for treatment of COPD-related ventilatory failure.
- Provided written, informed consent.

**Exclusion criteria**

- Diagnosis of neuromuscular disorders or kyphoscoliosis.
- Patients who, in the opinion of the investigator, are unlikely to comply with the requirements of the study, use the device correctly, or keep the diary records.

### General Breathing Record Study (GBRS)

**Primary Objective**

To explore the characteristics of the Tidal Breathing carbon dioxide ($TBCO_2$) waveform that can differentiate between different respiratory and cardiac conditions (including acute and chronic disease states) and establish a profile for healthy controls.

**Secondary Objectives**

- To identify within-patient changes in the $TBCO_2$ waveform that may predict or detect a deterioration of the underlying disease.
- To establish whether the characteristics of any waveform changes before a deterioration are similar in all patients in the same group.
- To identify waveform features that may help inform a larger disease-specific prospective study.
- To describe the relationship between characteristics of the $TBCO_2$ waveform and severity of the primary condition of interest (as measured by disease-specific clinical parameters and symptom questionnaires).
- To compare the use of the $TBCO_2$ waveform (and the N-Tidal C device) in monitoring different breathing conditions to traditional methods of monitoring disease control.
- To identify the correlation of carbon dioxide ($CO_2$) measurements by capnography with those obtained by periodic standard arterial blood gas measurements in patients with respiratory diagnoses.
- To monitor the safety of the device in regular use by patients at home.
- To establish the 'ease of use' of capnography measurement as a potential disease diagnosis and monitoring method in all participants.
- To establish the 'ease of use' of capnography measurement in HCP's.
- To identify the ranges of $TBCO_2$ waveform parameter values (minimum and maximum) for the different disease cohorts.
- To identify within-day and day-to-day variations in $TBCO_2$ waveform parameter values for the different disease cohorts.

**General Inclusion Criteria**

- Male or female, aged $\geq$ 16 years
- Willing and able to provide written informed consent.

**General Exclusion Criteria**

- Known other lung, chest wall, neuromuscular, cardiac or other comorbidity or abnormality that would affect spirometry and/or other measures of lung function or $TBCO_2$ measurements.
- In the opinion of the clinical investigator, the participant would have difficulty completing the study procedures consistently over 6 months.

**Asthma Inclusion Criteria**

- A confirmed clinical diagnosis of asthma for $\geq$ 6 months supported by evidence of any of the following:
  - Airflow variability, with a variability in FEV1 of >20% across clinic visits within the preceding 12 months, with concomitant evidence of airflow obstruction (FEV1/FVC ratio <70% on spirometry).
  - Airway reversibility with an improvement in FEV1 by $\geq$ 12% or 200 ml after inhalation of 400 $\mu$g of salbutamol via a metered dose inhaler and spacer within the preceding 12 months.
  - Airway hyper-responsiveness demonstrated by Methacholine (or similar) challenge testing with a provocative concentration of Methacholine required to cause a 20% reduction in FEV1 (PC20) of $\leq$ 8mg/ml or equivalent test.
- Moderate to severe asthma defined as BTS stage 3 5
- Exacerbation free for >2 weeks (defined as no increased dose or course of oral corticosteroids or antibiotics).
- 2 or more exacerbations in the previous 12 months with at least 1 exacerbation within the last 6 months.

**Breathing Pattern Disorder / Vocal Cord Dysfunction Inclusion Criteria**
A Clinical diagnosis of a Breathing Pattern Disorder (BPD) by a Specialist Respiratory Physiotherapist.

**Chronic Heart Failure**

- A confirmed clinical diagnosis of chronic heart failure with both of the following:
  - A Left Ventricular Ejection Fraction <40% on most recent imaging within the last 12 months.
  - New York Heart Association Class 2-4
- Either (i) admitted with an acute decompensation of their heart failure to hospital requiring intravenous diuretics or an increase in diuretic dose from baseline (e.g., 40mg or more furosemide) within the last 6 months or (ii) stable outpatient with NT-proBNP >400ng/mL in sinus rhythm or NT-proBNP >1000ng/mL in atrial fibrillation.

**Motor Neurone Disease**
A confirmed clinical diagnosis of Motor Neuron Disease (MND)

**Pneumonia**
A confirmed clinical diagnosis of Pneumonia supported by evidence of consolidation on a chest X-ray (CXR) or computed tomography (CT) imaging.

**Healthy Volunteers**

- No known history of lung, cardiac or neuromuscular disease (defined as no current clinical diagnosis of, or receiving treatment for, a lung, cardiac or neuromuscular disease).
- BMI $\leq$ 40
- Non-smoker, or ex-smoker with $\leq$ 5 pack year history

### COPD Breathing Record Study 2 (CBRS2)

**Primary Objective**
To assess the changes in key parameters in the tidal breathing $CO_2$ ($TBCO_2$) waveform, including the $\alpha$ angle, the minimum $CO_2$ level achieved and the stability of the expiratory cycle, during the transition from stable COPD to during acute exacerbations.

**Secondary Objectives**

- To collect a longitudinal observational database of $TBCO_2$ waveform records for up to 50 patients with moderate-to-severe COPD over 26 weeks using the N-Tidal C Data Collector Device.
- To capture the carbon dioxide signature predictive of exacerbations in monitored patients who undergo mild, moderate and severe exacerbations during the study.
- To identify the within-day and day-to-day variability of respired $CO_2$ in patients with COPD.

**Inclusion Criteria**

- Aged 40 years and over.
- Diagnosis of COPD (Primary) and at least one moderate exacerbation within 12 months of starting the study period.
- Able to provide signed informed consent.

**Exclusion Criteria**

- Patients who, in the opinion of the investigator, are unlikely to comply with the requirements of the study, use the device correctly or keep the diary records.
- Diagnosis of neuromuscular disorders or Kyphoscoliosis.
- Diagnosis of other Respiratory disorders that, in the investigator's opinion, would impact the conduct of the study e.g., clinically significant bronchiectasis, asthma.
- Patients who have experienced an exacerbation of their COPD that has required treatment with antibiotics and/or oral corticosteroids within 2 weeks before the study start.

#### Asthma Breathing Record Study (ABRS)

**Primary Objective**

To determine characteristics within the $TBCO_2$ waveform shape, as measured by the N-Tidal C data collector device, that identify deteriorations in the user's respiratory condition, including changes leading to asthma exacerbations, and discriminate between poorly and well-controlled asthma.

**Secondary Objectives**

- To determine whether changes in the $TBCO_2$ waveform, measured by the N-Tidal C data collector device, can predict asthma exacerbations.
- To describe the relationship between characteristics of the $TBCO_2$ waveform and severity of asthma at baseline, as measured by:
  - BTS Stage 2-5
  - Disease control (Asthma Control Questionnaire)
  - Quality of Life (Asthma Quality of Life Questionnaire)
  - Spirometry (% predicted FEV1)
  - Fractional exhaled Nitric Oxide (FeNO in ppb, if available)
  - Airway resistance (Airway Oscillometry, if available)
  - Peak Expiratory Flow (PEF)
  - Forced Expiratory Flow (FEF25-75)
- To analyze subgroups of exacerbations according to the presence of triggers.
- To explore the feasibility of monitoring asthma control at home using the N-Tidal C in all participants.
- To assess the usability and acceptability of the device to study participants and their family/carers, gathering ideas for improved use and further development.
- To assess the adherence to use of the device by participants and explore barriers and facilitators to adherence.
- To evaluate healthcare resource use, costs and quality-of-life measures over the study period.
- To describe and quantify any adverse device effects.

**Inclusion Criteria**

- Male or Female, aged $\geq$ 7 years.
- Confirmed clinician diagnosis of asthma by examination of medical records and based on accepted national and/or international criteria e.g., BTS/SIGN, or GINA
- Moderate or Severe asthma (defined as BTS stage 2-5)
- Poorly controlled asthma (defined as an ACQ score of $\geq$ 1)
- Exacerbation prone asthma (defined as at least 1 asthma exacerbation requiring oral corticosteroid treatment in the last 12 months)
- Capable of providing written informed consent, or parental/guardian consent and participant assent in the case of a child

**Exclusion Criteria**

- Inability to understand or comply with study procedures and/or give fully informed consent.
- Known other lung, chest wall, neuromuscular, cardiac or other comorbidity or abnormality that would affect spirometry and/or other measures of lung function or $TBCO_2$ measurements (including Breathing Pattern Disorder or Chronic Obstructive Pulmonary Disease).
- Smokers (current or ex-smokers) with a >10 pack year history.
- In the opinion of the clinical investigator, the participant would have difficulty completing the study procedures consistently (for example, difficulty holding the device, or long periods of absence/travel) throughout the study period.

| | **COPD Breathing Record Study (CBRS)** | **General Breathing Record Study (GBRS)** | **COPD Breathing Record Study 2 (CBRS2)** | **Asthma Breathing Record Study (ABRS)** |
|---|---|---|---|---|
| **Primary Objective** | To collect a longitudinal observation study database of capnograph records for up to 30 patients with recent or recurrent exacerbations of COPD over 6 weeks using. | To explore the characteristics of the $TBCO_2$ waveform that can differentiate between different respiratory and cardiac conditions (including acute and chronic disease states) and establish a profile for healthy controls. | To assess the changes in key parameters in the $TBCO_2$ waveform, including the $\alpha$ angle, the minimum $CO_2$ level achieved and the stability of the expiratory cycle, during the transition from stable COPD to during acute exacerbations. | To determine characteristics within the $TBCO_2$ waveform shape that identify deteriorations in the user's respiratory condition, including changes leading to asthma exacerbations, and discriminate between poorly and well-controlled asthma. |
| **Population** | Community group - chronically elevated $CO_2$ and frequent exacerbations of COPD; Acute admissions group – admitted to hospital with an exacerbation of COPD. | Disease cohorts selected from hospital outpatient clinic lists and inpatient wards. Healthy volunteers. | Patients with moderate-to-severe COPD from the Cambridge COPD center. | Patients with poorly controlled asthma. |
| **Clinical condition(s)** | COPD | Asthma, breathing pattern disorder, chronic heart failure, motor neuron disease, pneumonia, healthy volunteers. | COPD | Asthma |
| **Number of participants** | COPD: 30 | Asthma: 20; BPD, CHF, MND, Pneumonia, Healthy 10 each; Total: 70 | COPD: 50 | Asthma: 124; 92 of these were recruited from primary care, and 32 from secondary care. |
| **Location** | Addenbrooke's Hospital, Cambridge University Hospitals NHS Foundation Trust, UK | Queen Alexandra Hospital and specialist secondary care community clinics, Portsmouth Hospitals University NHS Trust, UK | COPD Centre, Addenbrooke's Hospital, Cambridge University Hospitals NHS Foundation Trust, UK | Queen Alexandra Hospital, Portsmouth Hospitals University NHS Trust, UK and GP practices, Oxford, UK |
| **Recruitment setting** | Outpatient, Inpatient | Outpatient | Outpatient | Outpatient, Inpatient and Primary Care |
| **Duration** | 17th Feb 2016 – Dec 2016 | 9th Aug 2017 – 4th Jul 2018 | 15th Aug 2017 – 23rd Nov 2018 | 11th Feb 2020 – 31st Jan 2022 |
| **Clinical Trials.gov Identifier** | NCT02814253 | NCT03356288 | NCT03615365 | NCT04504838 |
| **Number of capnograms** | 2620 | 15803 | 14885 | 38026 |
| **Additional data collected** | Medical history, clinical assessment, demographics, vital signs, spirometry, routine blood tests, blood gases. | Medical history, clinical assessment, demographics, vital signs, spirometry (not heart failure or pneumonia groups). Other assessments were disease specific. | Medical history, clinical assessment, demographics, symptoms, vital signs, spirometry. | Medical history, clinical assessment, demographics, vital signs, spirometry (plus optional others - FeNO, oscillometry, full body plethysmography) routine blood tests. |

**Table 1.** Summary of the four clinical studies from which the paper has drawn its data.
